# The Burden of Gallstone Disease in the United States Population

**DOI:** 10.1101/2022.07.08.22277386

**Authors:** Aynur Unalp-Arida, Constance E. Ruhl

## Abstract

**Background and rationale:** Gallstone disease is one of the most common digestive disorders in the United States and leads to significant morbidity, mortality, and health care utilization. We used national survey and claims databases to expand on earlier findings and investigate current trends in the gallstone disease burden in the United States.

**Methods:** The National Ambulatory Medical Care Survey, National Inpatient Sample, Nationwide Emergency Department Sample, Nationwide Ambulatory Surgery Sample, Vital Statistics of the U.S., Optum Clinformatics® Data Mart, and Centers for Medicare and Medicaid Services Medicare 5% Sample databases were used to estimate claims-based prevalence, medical care including cholecystectomy, and mortality with a primary or other gallstone diagnosis. Rates were age-adjusted (for national databases) and shown per 100,000 population.

**Results:** Gallstone disease prevalence (claims-based, 2019) was 0.72% among commercial insurance enrollees and 2.09% among Medicare beneficiaries and rose over the previous decade in both groups. Recently, in the U.S. population, gallstone disease contributed to approximately 2.2 million ambulatory care visits, 1.2 million emergency department visits, 625,000 hospital discharges, and 2,000 deaths annually. Women had higher medical care rates with a gallstone disease diagnosis, but mortality rates were higher among men. Hispanics had higher ambulatory care visit and hospital discharge rates compared with Whites, but not mortality rates. Blacks had lower ambulatory care visit and mortality rates, but similar hospital discharge rates compared with whites. During the study period, ambulatory care and emergency department visit rates with a gallstone disease diagnosis rose, while hospital discharge and mortality rates declined. Among commercial insurance enrollees, rates were higher compared with national data for ambulatory care visits and hospitalizations, but lower for emergency department visits. Cholecystectomies performed in the U.S. included 605,000 ambulatory laparoscopic, 280,000 inpatient laparoscopic, and 49,000 inpatient open procedures annually. Among commercial insurance enrollees, rates were higher compared with national data for laparoscopic procedures

**Conclusion:** The gallstone disease burden in the United States is substantial and increasing, particularly among women, Hispanics, and older adults with laparoscopic cholecystectomy as the mainstay treatment. Current practice patterns should be monitored for better health care access.

## INTRODUCTION

Gallstones (cholelithiasis) are concretions, usually composed of cholesterol or bilirubin, which develop in the gallbladder. Gallstones may form if bile contains excess cholesterol or bilirubin or insufficient bile salts. Most gallstones in the United States and western countries are cholesterol gallstones. Risk factors include older age, female sex, Native American and Hispanic ethnicity, overweight or obesity, diabetes, smoking, higher parity among women, rapid weight loss, and genetic factors. Non-Hispanic black race-ethnicity, greater alcohol consumption, increased serum cholesterol, and greater physical activity are associated with lower risk.(1) Though the majority remain asymptomatic, gallstones can block bile ducts causing acute right upper quadrant pain and if untreated, can lead to complications, including choledocholithiasis, cholangitis, and cholecystitis. Symptomatic gallstones are usually treated by cholecystectomy.

Gallstones are common and lead to significant morbidity, mortality, and health care utilization in the United States and worldwide. More than 20 million people in the United States have ultrasound-detected gallbladder disease.(1) There were an estimated 1.8 million ambulatory care visits with an all-listed gallstone diagnosis in 2004 and rates were relatively stable over time.(2) There were 622,000 overnight hospitalizations with an all-listed gallstone diagnosis in 2004. Hospitalization rates declined by 40% from 1991 due to the shift to outpatient laparoscopic cholecystectomy.(2) There were 2,155 deaths with gallstones as underlying or other cause in 2004 and mortality rates fell between 1979 and 2004 by 70%.(2,3) Based on data from the National Survey of Ambulatory Surgery conducted by the National Center for Health Statistics of the Centers for Disease Control and Prevention, gallstone disease resulted in 503,000 laparoscopic cholecystectomies in 2006.(4) A more recent report on the burden of gastrointestinal, liver, and pancreatic disease in the U.S. found that cholelithiasis was the physician diagnosis for 863,000 office visits and 327,000 emergency department visits in the U.S. in 2016.(5) Cholelithiasis and cholecystitis was the 10^th^ most common among all-listed digestive disease diagnoses from emergency department visits in 2018 with 1.5 million visits and the 5th most common among all-listed digestive disease diagnoses in U.S. hospitals in 2018 with 741,000 admissions.(5)

Current estimates of the gallstone disease burden in the United States are useful to all multidisciplinary clinicians, researchers, public health professionals, and policy makers for better planning. We used national survey and claims databases to expand on earlier findings and estimate current trends in the gallstone disease burden in the United States.

## MATERIALS AND METHODS

### Data sources

The National Ambulatory Medical Care Survey (NAMCS), Healthcare Cost and Utilization Project (HCUP) National Inpatient Sample (NIS), HCUP Nationwide Emergency Department Sample (NEDS), HCUP Nationwide Ambulatory Surgery Sample (NASS), Vital Statistics of the U.S., Optum Clinformatics® Data Mart (CDM), and the Centers for Medicare and Medicaid Services (CMS) Medicare 5% Sample databases were used to estimate claims-based prevalence, medical care including cholecystectomy, and mortality with a primary or other gallstone diagnosis.

#### National databases

The NAMCS is conducted in the United States by the National Center for Health Statistics of the Centers for Disease Control and Prevention (CDC).(6) It is an annual nationally representative sample survey of office-based outpatient visits to non-federal health care providers. It is a multistage stratified probability sample of geographically defined areas, physician practices within these areas, and patient visits within physician practices. In 2006, a separate sample of community health centers was added to the survey. Providers were asked to complete patient encounter forms for a systematic random sample of visits occurring during a randomly assigned one-week reporting period, including the first four provider diagnoses.

The HCUP NIS, NEDS, and NASS are maintained by the Agency for Healthcare Research and Quality. The HCUP NEDS consists of a nationally representative sample of hospital-owned emergency department visits from non-federal hospitals collected annually regardless of whether they result in admission.(8) The 2020 NEDS includes discharge data for emergency department visits from 995 hospitals located in 40 States and the District of Columbia, approximating a 20-percent stratified sample of U.S. hospital-owned emergency departments. It contains data from over 30 million emergency department visits each year (unweighted) and estimates approximately 145 million emergency department visits (weighted).

The NIS consists of a nationally representative sample of discharges from all participating non-federal hospitals collected annually.(7) The 2020 NIS was drawn from 48 participating states and the District of Columbia covering more than 97% of the U.S. population and approximates a 20% stratified sample of all discharges from U.S. community hospitals, excluding rehabilitation and long-term acute care hospitals. It contains data from more than 7 million hospital stays each year (unweighted) and estimates more than 35 million hospitalizations nationally (weighted). Data collected include up to 15 diagnoses and 7 surgical procedures.

The HCUP NASS consists of a nationally representative sample of major ambulatory surgery encounters performed in hospital-owned facilities.(9) The 2020 NASS includes data from 2,899 hospital-owned facilities located in 35 states and the District of Columbia, approximating a 67-percent stratified sample of U.S. hospital-owned facilities performing selected ambulatory surgeries. It contains approximately 7.8 million ambulatory surgery encounters each year and approximately 10.8 million ambulatory surgery procedures (unweighted) and estimates approximately 10.3 million ambulatory surgery encounters and 13.5 million ambulatory surgery procedures (weighted). Major ambulatory surgeries are defined as selected major therapeutic procedures that require the use of an operating room, penetrate or break the skin, and involve regional anesthesia, general anesthesia, or sedation to control pain.

The Vital Statistics of the U.S.: Multiple Cause-of-Death Data are managed by CDC and include all deaths occurring within the United States collected annually.(10) An underlying cause and up to 20 contributing causes of death were derived from death certificates.

#### Claims databases

Optum’s CDM is comprised of administrative health care claims for members of large commercial health plans and since 2006, Medicare Advantage with Part D health plans.(11) Administrative claims submitted by providers and pharmacies are verified, adjudicated, and de-identified before inclusion in CDM. Data are included for members with both medical and prescription drug coverage. The population includes individuals in all 50 states. The database includes approximately 17 to 19 million annual covered individuals for over 68 million unique members from January 2007 through December 2020.

The Medicare 5% sample was created by CMS to be representative of the entire population of Medicare beneficiaries and is housed in the CMS Chronic Conditions Data Warehouse.(12) Data are linked by a unique, unidentifiable beneficiary key. The Medicare datasets contain fee-for-service and Advantage institutional and non-institutional claims, enrollment/eligibility information (Part A, Part B, Part D and Medicare Advantage (Part C)), and NDI Cause of Death data.

### Gallstone disease and cholecystectomy definitions

Gallstone disease morbidity was identified by an International Classification of Diseases (ICD), Ninth Revision, Clinical Modification (ICD-9-CM) code of 574 or an ICD, Tenth Revision, Clinical Modification (ICD-10-CM) code of K80, and gallstone disease mortality by an ICD, Tenth Revision (ICD-10) code of K80. The first-listed diagnosis was considered the primary diagnosis and all remaining diagnoses were considered secondary and included under ‘all-listed’. For national event-level data sources, diagnoses were counted only once under the all-listed category, irrespective of the number of actual diagnoses listed on a medical record or death certificate. In other words, if there was more than one gallstone disease diagnosis listed on a medical record or death certificate, only one diagnosis was counted for the ‘all-listed’ category. Cholecystectomy procedure codes are shown below.

| Type | Current Procedural Terminology (CPT®) | ICD-9-PCS | ICD-10-PCS |
| --- | --- | --- | --- |
| Laparoscopic | 47562, 47563, 47564 | 51.23, 51.24 | 0FT44, 0FB44 |
| Open | 47600, 47605, 47610, 47612, 47620 | 51.21, 51.22 | 0FB40, 0FB43<br>0FT40 |

### Statistical analysis

Because of potential short-term effects of the COVID-19 pandemic on gallstone disease healthcare utilization and mortality, statistics highlighted in the text are for 2019, while figures present trends through 2020.

#### National databases

Estimates for the total population and by age, sex, race, and Hispanic origin were calculated for each year of national data. Event counts are shown as number in thousands. For calculating U.S. population rates, annual population data were derived from the national population estimates program of the U.S. Census Bureau and the CDC. For all rate calculations, population estimates used as the denominator were mid-year population counts by age, sex, race, and Hispanic origin. Rates were age-adjusted by direct standardization using the year 2000 population estimates and are shown per 100,000 population.(13) NAMCS, NEDS, NIS, and NASS data were weighted to generate national estimates. Ambulatory care visit (NAMCS) data points were 3-year averages to provide more stable estimates. Ambulatory care numbers and rates represent visits, not persons with a visit. Hospital numbers and rates represent discharges, not persons with an inpatient stay.

#### Claims databases

Enrollees/beneficiaries were considered gallstone disease patients if they had one or more claims with an ICD-9-CM or ICD-10-CM diagnostic code indicative of gallstone disease in any diagnostic code field. The claims-based prevalence was calculated as the percentage of privately insured enrollees/beneficiaries who qualified as gallstone disease patients each year. Estimates were reported overall and stratified by age, gender, race/ethnicity, and region.

Optum© CDM member eligibility files containing birth year, demographics, and eligibility period were linked to inpatient confinement (acute care hospital or skilled nursing facility stay), medical (health care professional services), medical diagnosis, and pharmacy claims files. All privately insured enrollees with a single consistent birth year recorded in CDM who resided in the U.S. and were continuously enrolled for at least one full calendar year were included. Inpatient hospitalizations were defined as having an inpatient hospital place of service. Ambulatory visits were evaluation & management visits with an office, outpatient hospital, or ambulatory surgical center place of service. Evaluation & management visits were additionally characterized by a non-facility location with a type of service including office visits, surgery, ER, consultations, or home health/hospice visits, and excluding non-encounter services.

The CMS Medicare 5% sample study population/denominator consisted of beneficiaries who were 65 years or older as of January 1st of the year, resided in the 50 U.S states or District of Columbia, and were enrolled in fee-for-service Part A or Part B Medicare benefits or in the Medicare Advantage program (Part C/Health Maintenance Organization benefits) at any time during the year. The denominator file which contains demographic and enrollment data was linked by the beneficiary unique identifier to institutional (hospital inpatient stays, hospital outpatient services, skilled nursing facilities, home health agencies and hospice care organizations) or non-institutional (also called Part B; health care professionals, supplies, and services) medical claims files. Estimates from the 5% sample are multiplied by 20 to represent national estimates.

## RESULTS

### Claims-based prevalence (Figures 2A-2C and Figures 3A-3B)

Among 13.7 million commercial insurance enrollees, 99,000 had a gallstone disease diagnosis resulting in a claims-based prevalence of 0.72% (2019). Claims-based prevalence was over twice as high among 54.4 million Medicare beneficiaries, among whom 1.1 million had a gallstone disease diagnosis for a claims-based prevalence of 2.09% (2019). Among commercial insurance enrollees, prevalence was higher among women compared with men and increased with age. It was highest among Hispanics, followed by Blacks, then Whites, and was lowest among Asians. In contrast, among Medicare beneficiaries, men had higher prevalence compared with women in recent years and Whites had higher prevalence compared with Blacks. Prevalence rose over time in both groups, by two-thirds (0.43% to 0.72% from 2007 to 2019) among commercial insurance enrollees and by almost a third (1.59% to 2.09% from 2006 to 2019) among Medicare beneficiaries. This increasing trend was seen among each sex and race-ethnicity group but was primarily limited to persons 65 years and older.

### Health care utilization - national data (Figures 1A-1I)

Gallstones contributed to 2.2 million ambulatory care visits (3-year average for 2014 to 2016), 1.2 million emergency department visits (2019), and 625,000 hospital discharges (2019). Ambulatory care visit, emergency department visit, and hospital discharge rates were all higher among women compared with men. Ambulatory care visit and hospital discharge rates were highest among Hispanics. Ambulatory care visit rates were lower among Blacks compared with Whites, but hospital discharge rates were similar. Ambulatory care visit rates increased with age. Emergency department visit rates were highest among older adults and similar among middle aged and younger adults. Hospital discharge rates increased with age, especially among persons 65+ years. Hospital discharge rates underestimate the actual burden because most hospitalizations with gallstones were for cholecystectomy and a high proportion of cholecystectomies were performed laparoscopically without an overnight stay, and therefore, were not included in hospitalization statistics.

**Figure 1.**
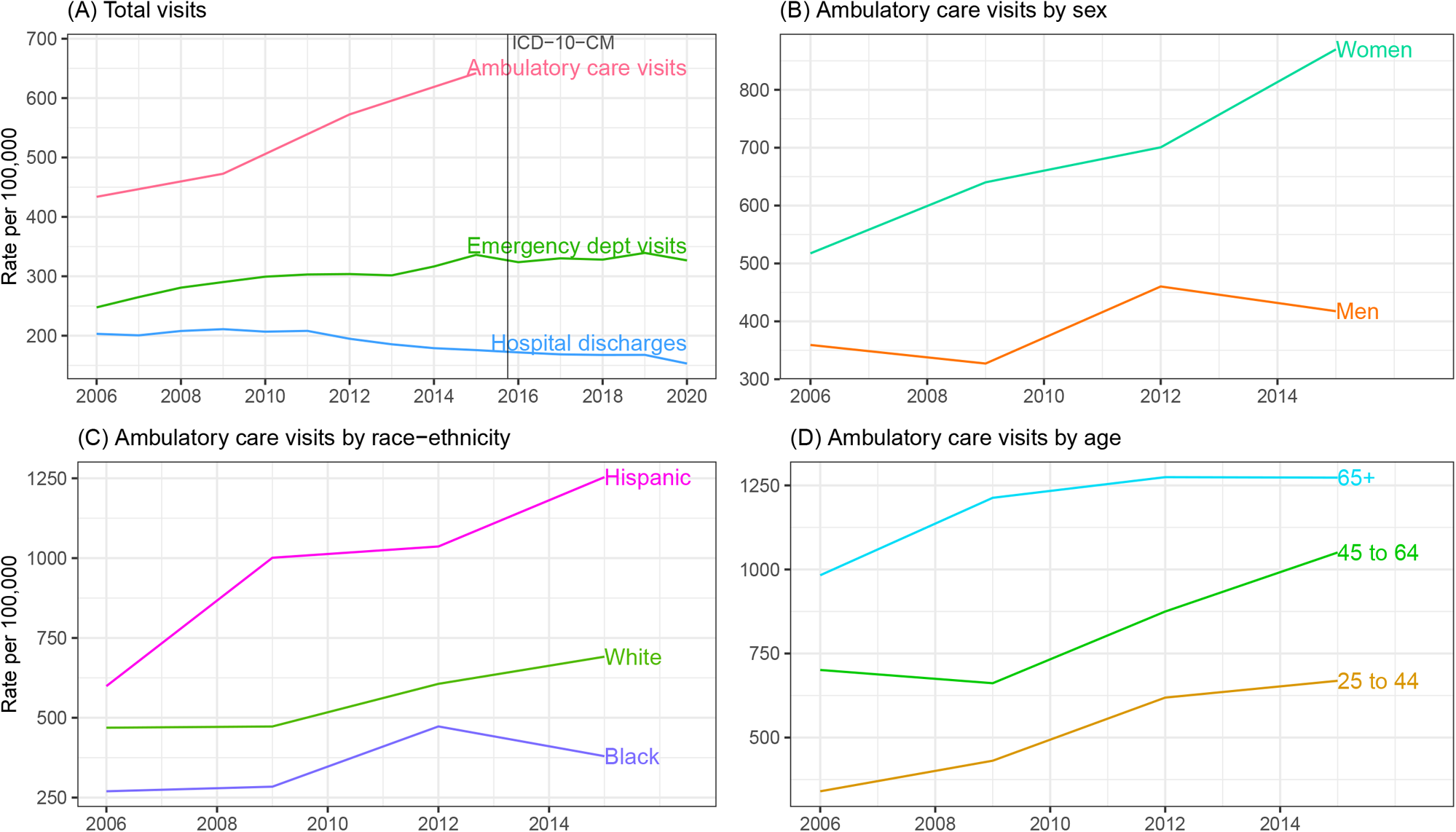
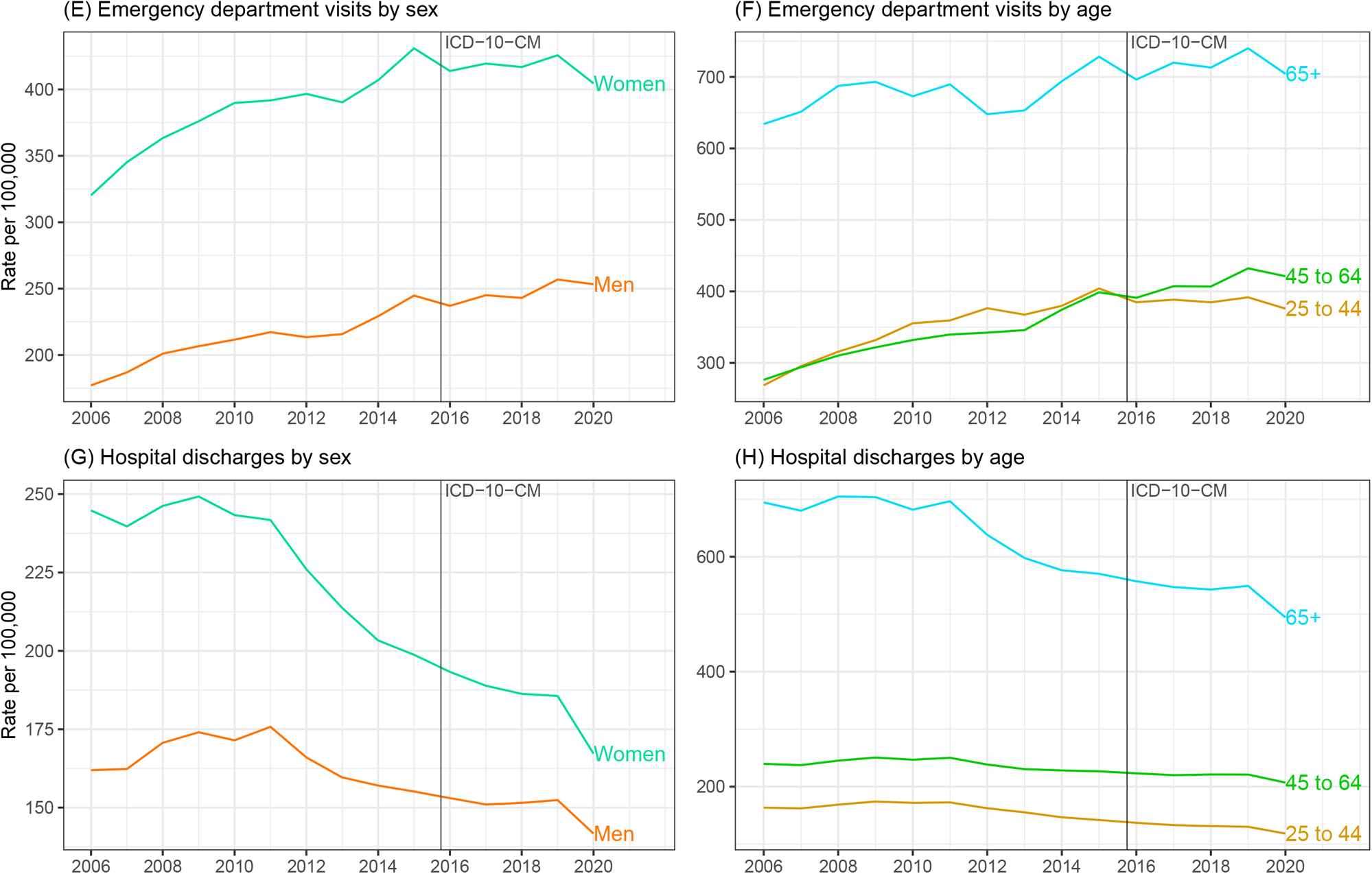
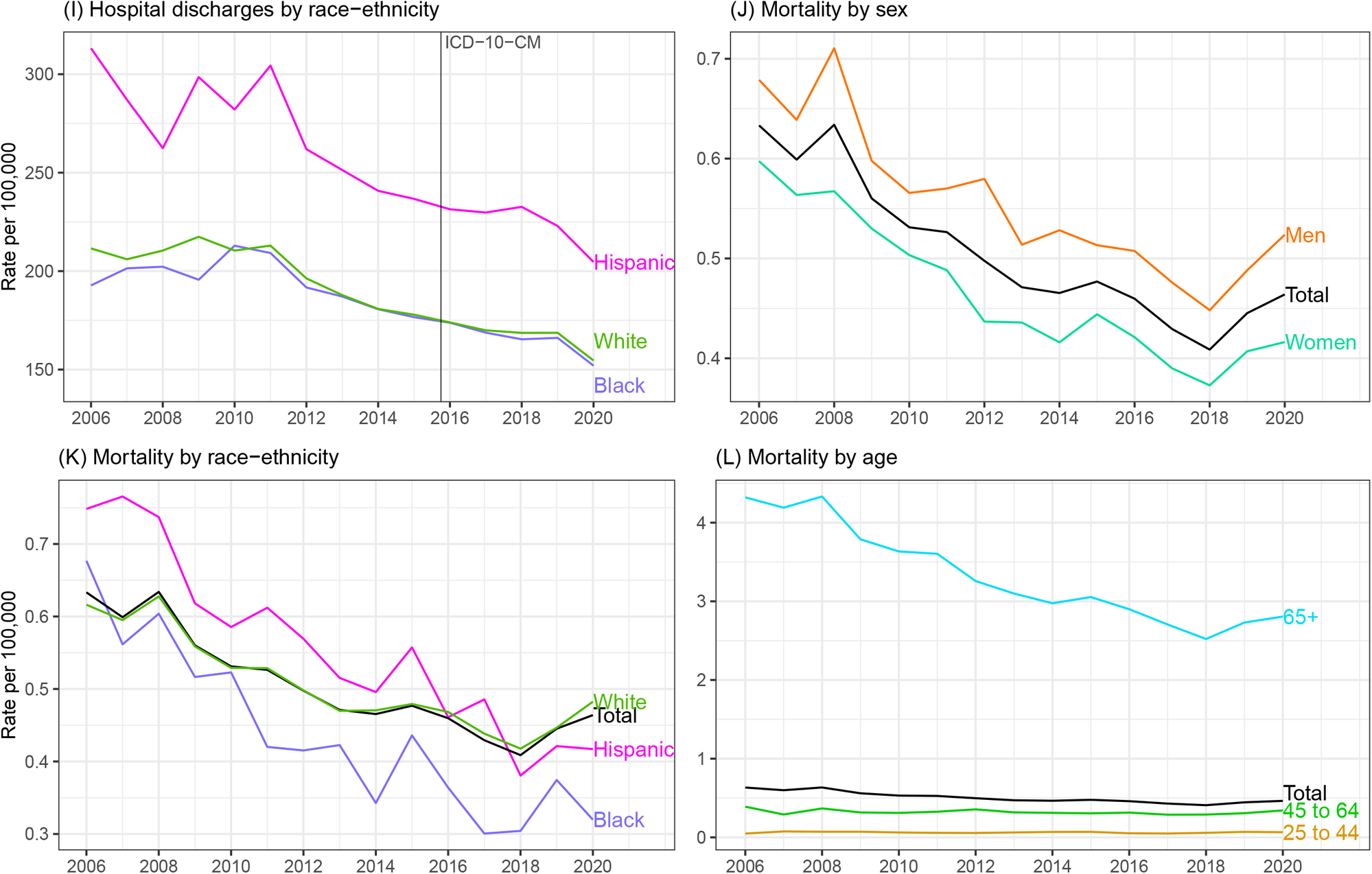
Age-adjusted rates with all-listed gallstone disease diagnoses in the United States (Source: National Ambulatory Medical Care Survey, 2005-2016 (3-year averages, 2005-2007, 2008-2010, 2011-2013, 2014-2016); Healthcare Cost and Utilization Project (HCUP) Nationwide Emergency Department Sample, 2006-2020; HCUP National Inpatient Sample, 2005-2020; and Vital Statistics of the United States, 2006-2020) A. Total visits B. Ambulatory care visits by sex C. Ambulatory care visits by race-ethnicity D. Ambulatory care visits by age E. Emergency department visits by sex F. Emergency department visits by age G. Hospital discharges by sex H. Hospital discharges by age I. Hospital discharges by race-ethnicity J. Mortality by sex K. Mortality by race-ethnicity L. Mortality by age

From 2005 to 2016, the ambulatory care visit rate (all-listed per 100,000) increased by almost 50% overall (434 to 642) and among sex, racial-ethnic, and age groups. From 2006 to 2019, the emergency department visit rate (all-listed per 100,000) increased by more than a third (248 to 339). A rise was seen in both women and men and among all age groups. In contrast, the hospital discharge rate (all-listed per 100,000) decreased by 17% (203 to 168) from 2006 to 2019. A decline was seen among each sex, race, and ethnic group, but primarily among older adults.

### Health care utilization - commercial insurance data (Figures 2D-2M)

Among commercial insurance enrollees, rates (all-listed per 100,000) were higher compared with national data for ambulatory care visits (799 vs. 642 in 2015) and hospitalizations (245 vs. 168 in 2019), but lower for emergency department visits (107 vs. 339 in 2019). Like national data, ambulatory care and emergency department visit rates were higher among women compared with men, but for hospitalization rates, the sex difference among commercial insurance enrollees has narrowed in recent years. Like national data, ambulatory care visit and hospital discharge rates were higher among Hispanics compared with Whites. In contrast to national data, Blacks had higher medical care rates compared with Whites among the commercially insured. Medical care rates were lowest for Asians. In contrast to national data in which hospitalization rates declined over the years studied, all three medical care rates increased among commercial insurance enrollees, primarily due to increases among persons 65 years and over.

**Figure 2.**
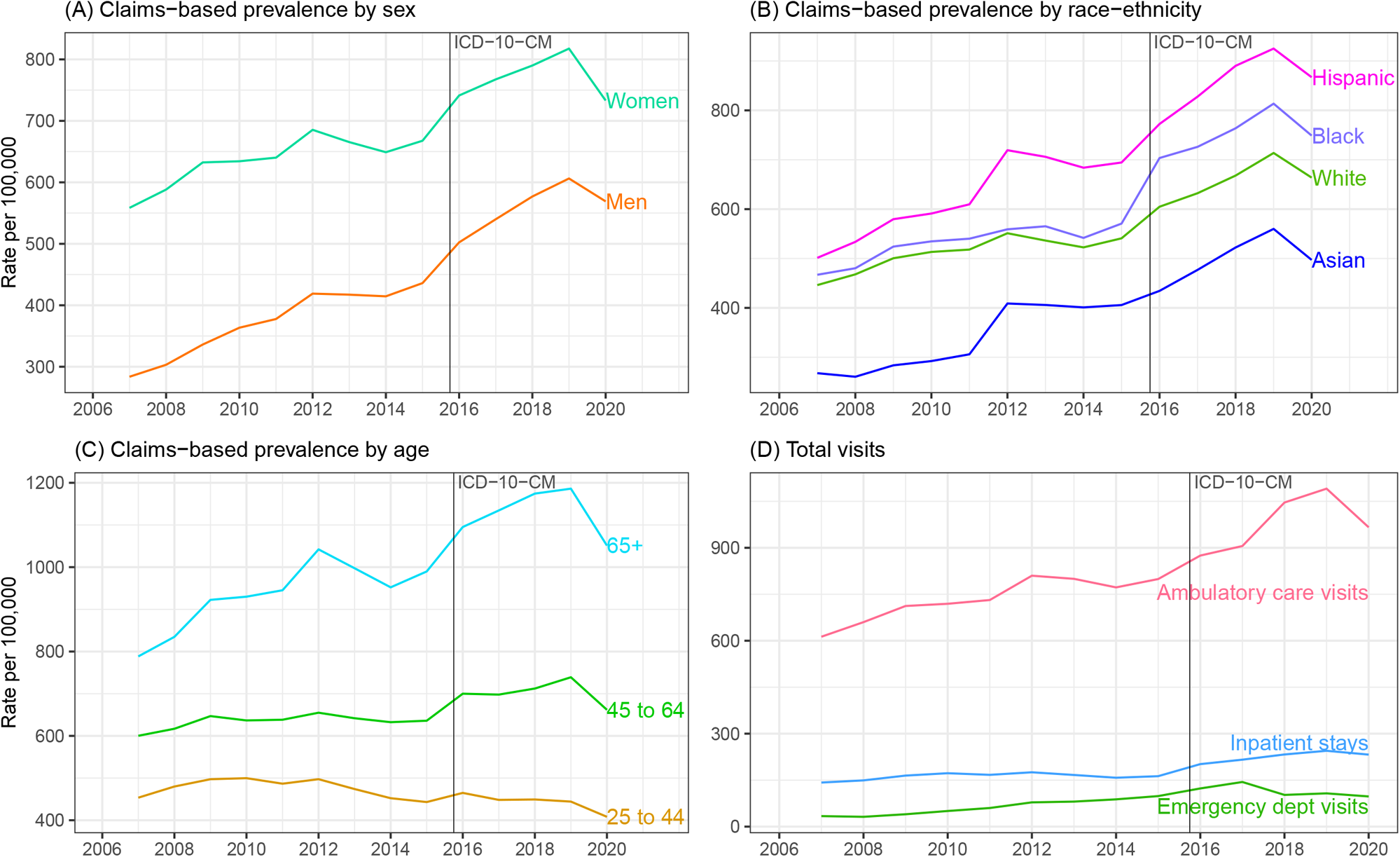
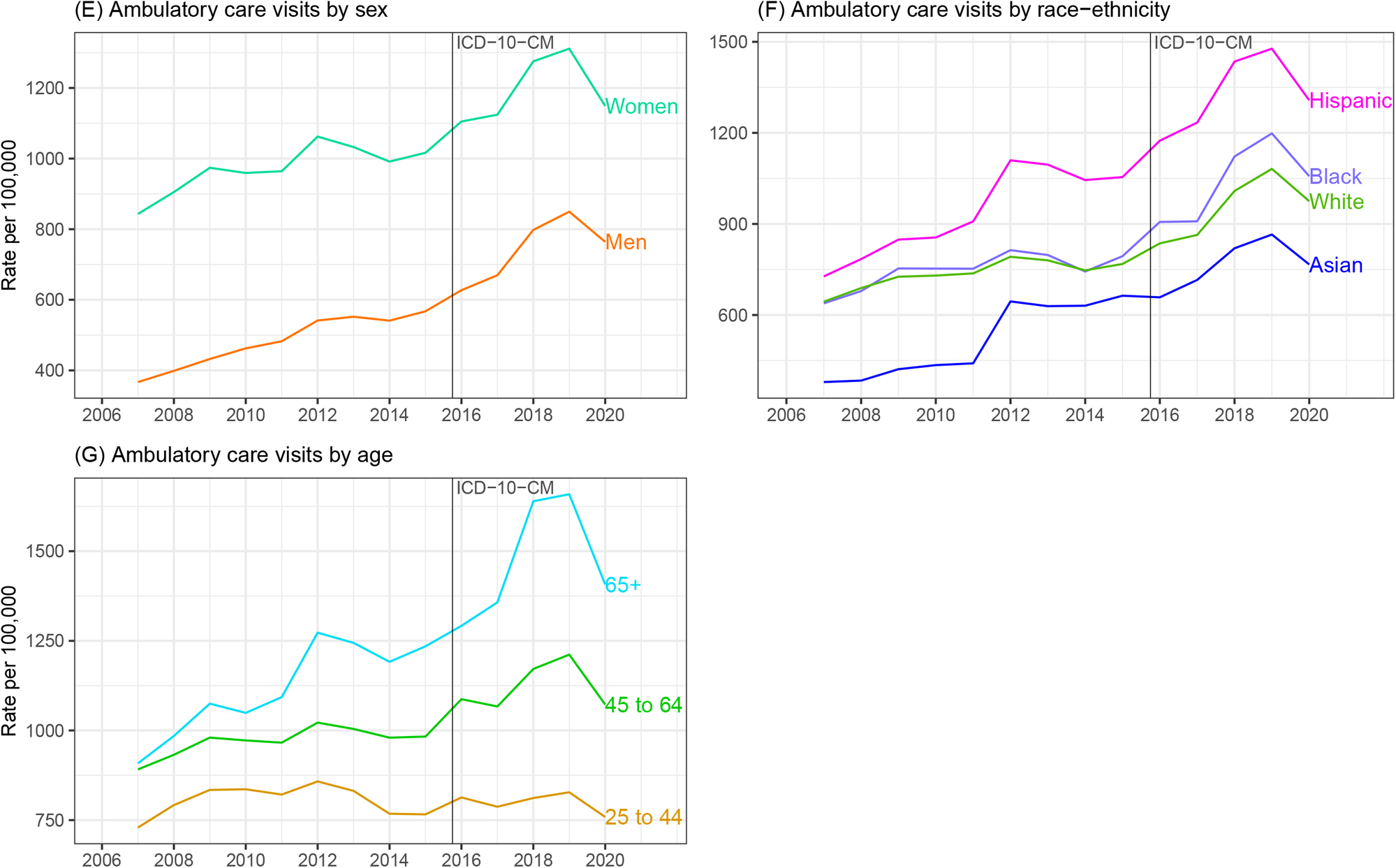
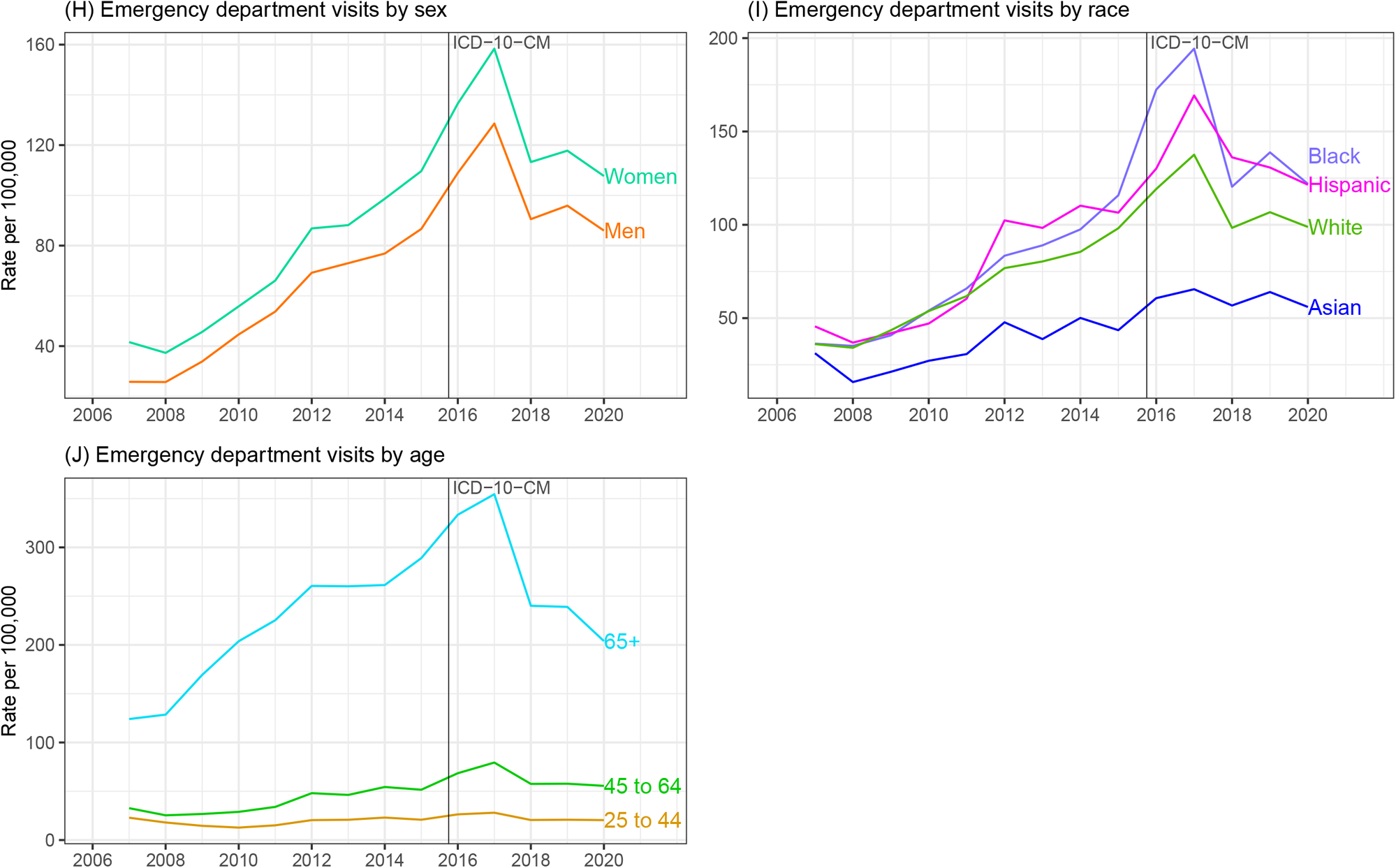
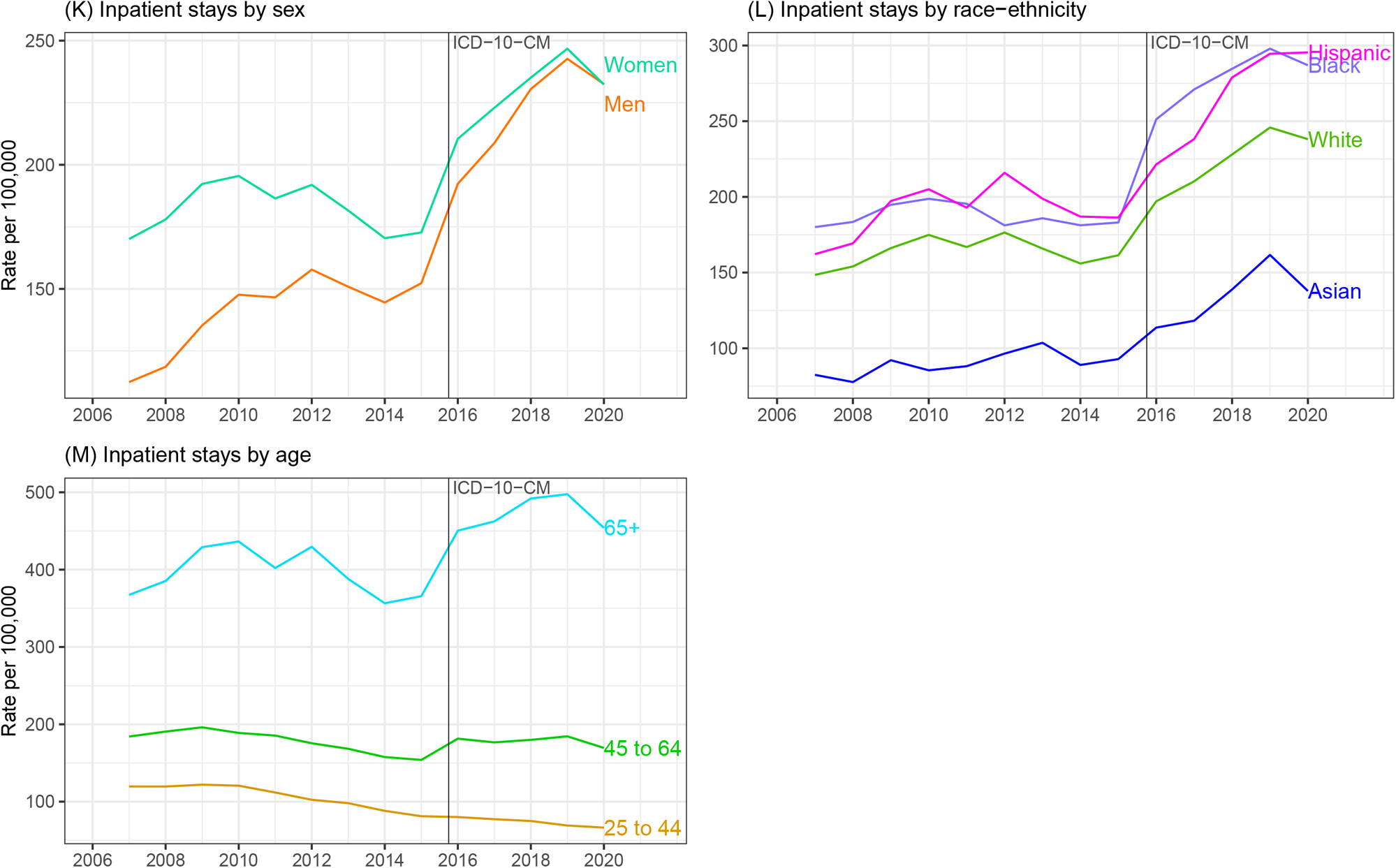
Rates with all-listed gallstone disease diagnoses among persons with commercial insurance in the United States, 2007-2020 (Source: Optum Clinformatics® Data Mart) A. Claims-based prevalence by sex B. Claims-based prevalence by race-ethnicity C. Claims-based prevalence by age D. Total visits E. Ambulatory care visits by sex F. Ambulatory care visits by race-ethnicity G. Ambulatory care visits by age H. Emergency department visits by sex I. Emergency department visits by race-ethnicity J. Emergency department visits by age K. Inpatient stays by sex L. Inpatient stays by race-ethnicity M. Inpatient stays by age

### Health care utilization - Medicare data (Figures 3C-3I)

Among Medicare beneficiaries, rates (all-listed per 100,000) were higher compared with national data for ambulatory care visits (2221 vs. 642 in 2015), emergency department visits (1134 vs. 339 in 2019) and hospitalizations (509 vs. 168 in 2019). Ambulatory care visit rates were higher among women, but in contrast to national data that included persons of all ages, emergency department visit and hospitalization rates were higher among male compared with female Medicare beneficiaries. Ambulatory care visit rates were lower among Blacks compared with Whites, but in contrast to national data, hospitalization rates were generally higher among Black compared with White Medicare beneficiaries. Emergency department visit rates were also higher among Blacks compared with Whites.

**Figure 3.**
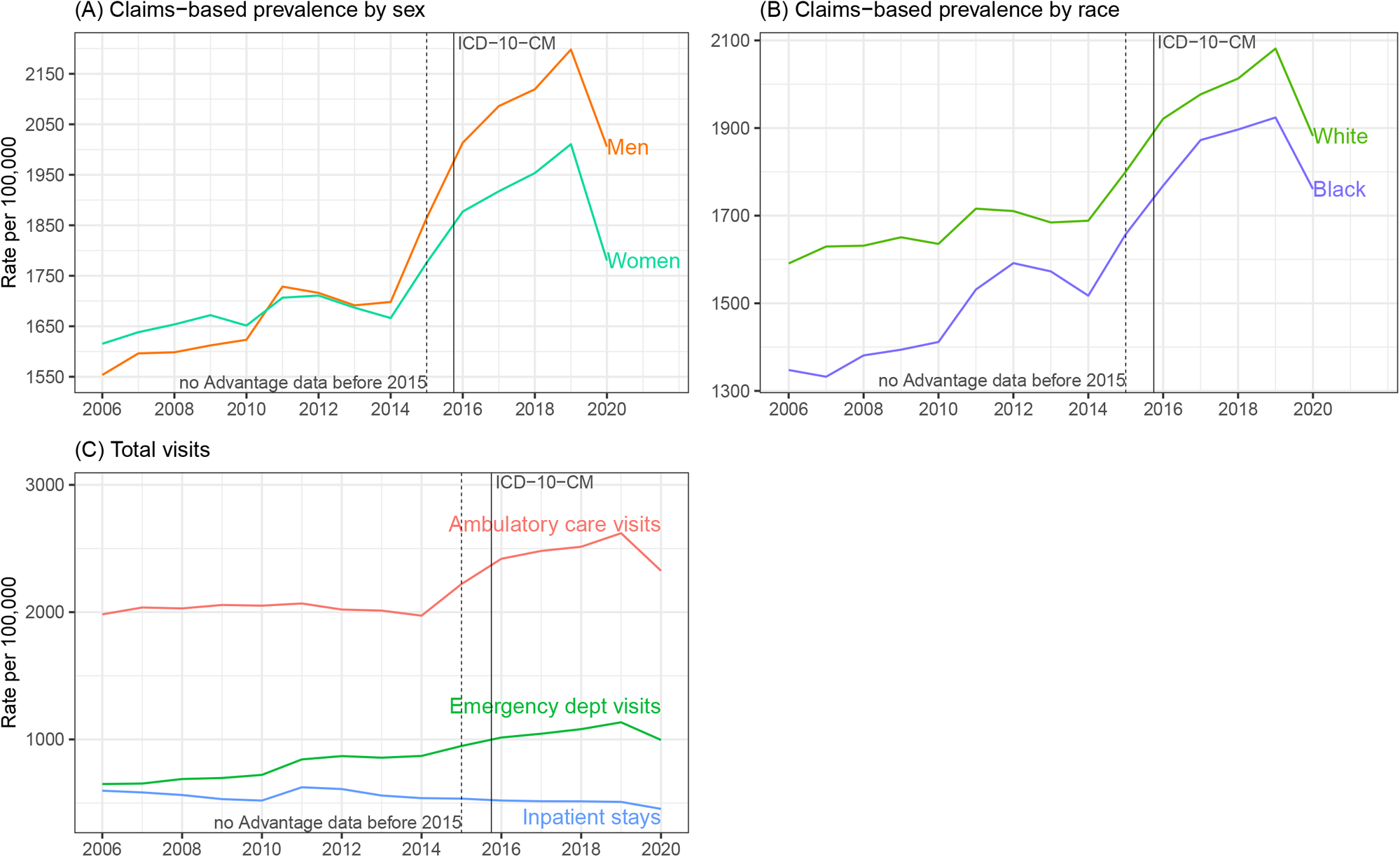
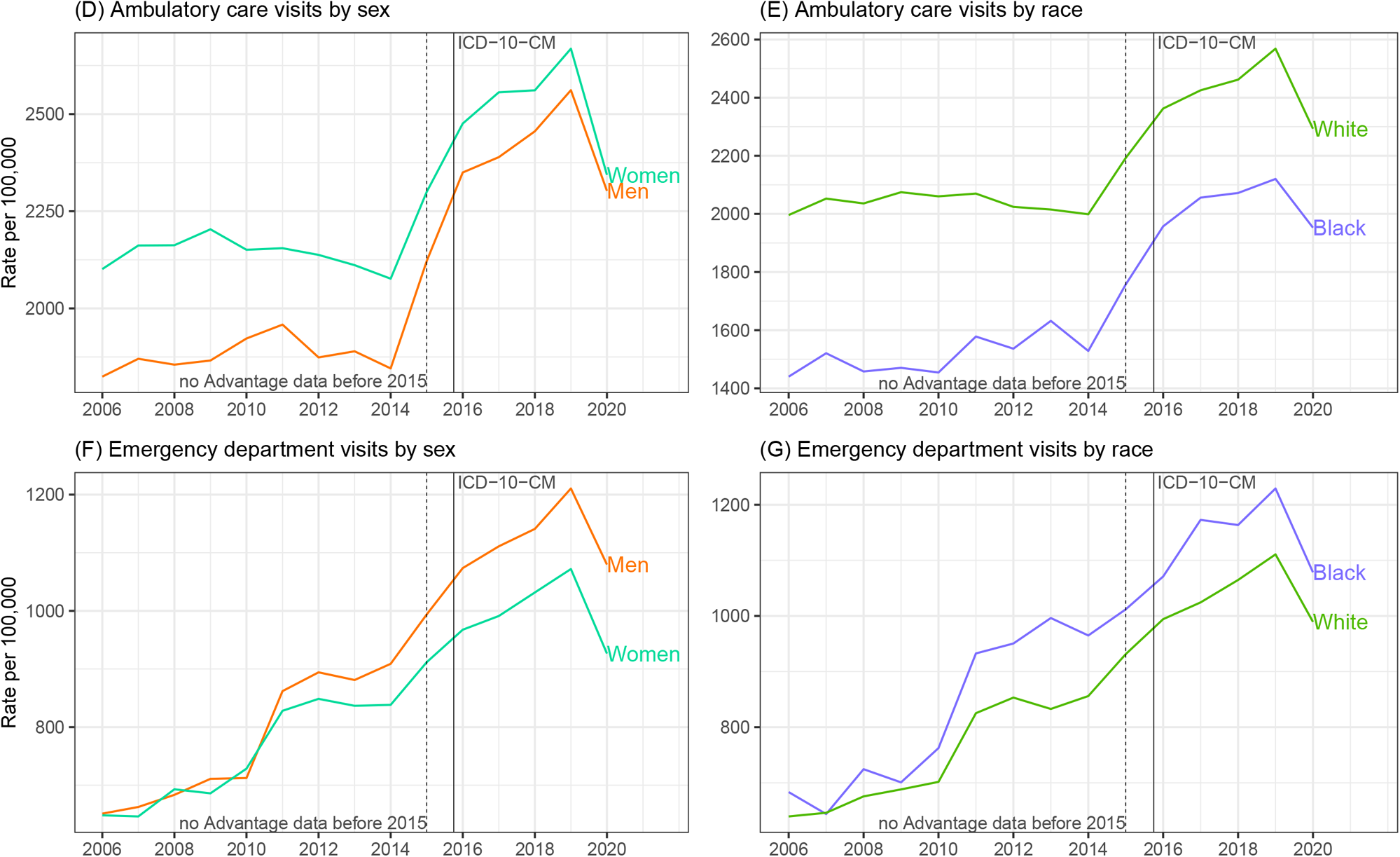
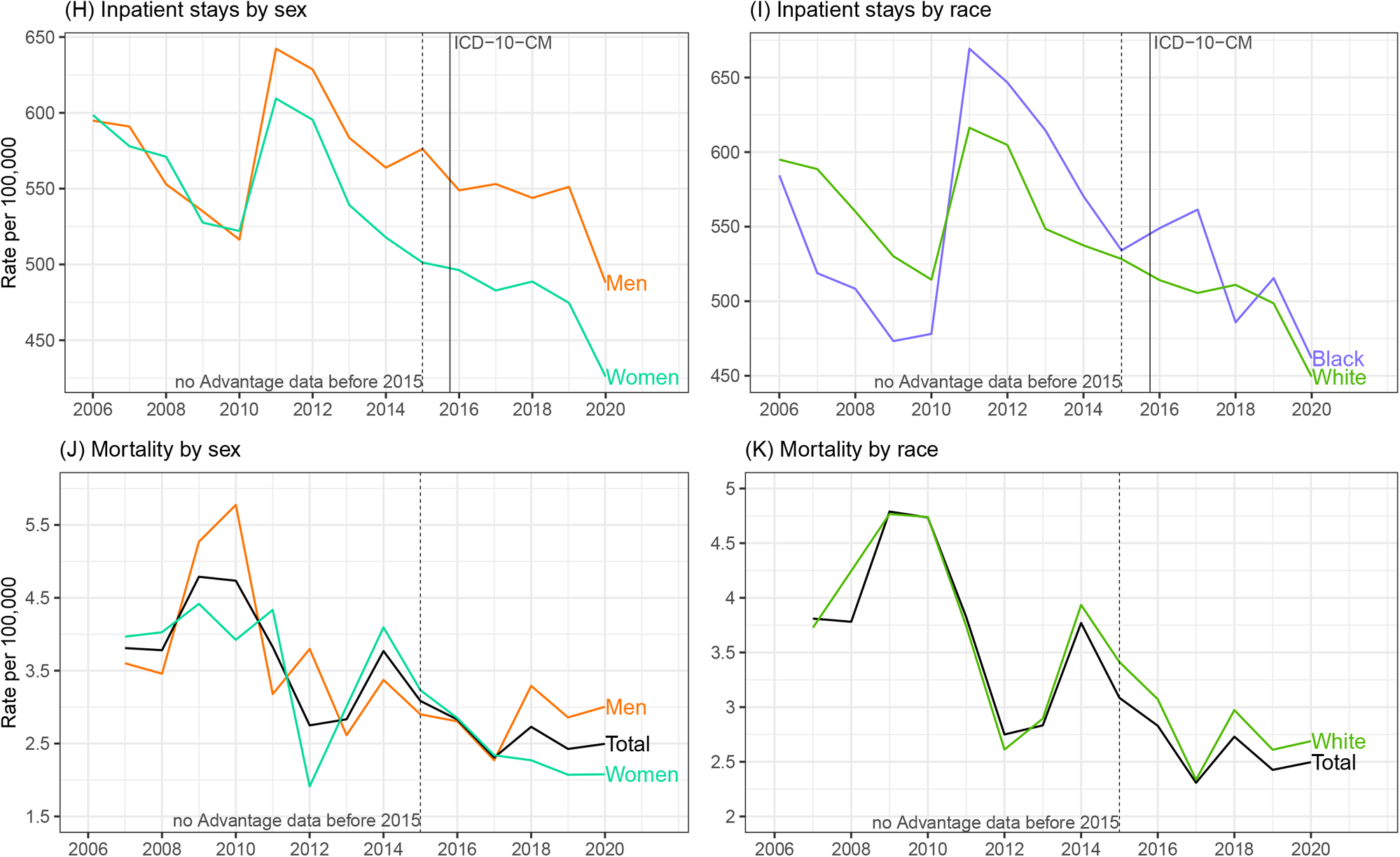
Rates with all-listed gallstone disease diagnoses among age-eligible fee-for-service Medicare beneficiaries in the United States, 2006-202) (Source: CMS Medicare 5% file) A. Claims-based prevalence by sex B. Claims-based prevalence by race C. Total visits D. Ambulatory care visits by sex E. Ambulatory care visits by race F. Emergency department visits by sex G. Emergency department visits by race H. Inpatient stays by sex I. Inpatient stays by race J. Mortality by sex K. Mortality by race

### Mortality (Figures 1J-1L and Figure 3J)

Gallstones contributed to 2,000 deaths in the U.S. (2019). Mortality rates were higher among men compared with women and much higher among persons 65+ years compared with younger adults. Rates were lower for Blacks compared with Whites and slightly lower for Hispanics compared with Whites after a steeper decline among Hispanics. From 2006 through 2019, the mortality rate (underlying or other cause per 100,000) decreased by more than a quarter overall (0.63 to 0.45) and among each sex and racial-ethnic group, but primarily among adults 65+ years.

### Cholecystectomy - national data (Figures 4A-4D)

In 2019, cholecystectomies performed in the U.S. included 605,000 ambulatory laparoscopic, 280,000 inpatient laparoscopic, 49,000 inpatient open, and 1,000 ambulatory open procedures. Rates (per 100,000) were highest for ambulatory laparoscopic (178), followed by inpatient laparoscopic (78), inpatient open (13), and ambulatory open (0.4) procedures. Laparoscopic cholecystectomy rates were higher among women compared with men. Racial-ethnic rates were not available for ambulatory laparoscopic cholecystectomies, but for inpatient laparoscopic procedures, rates were higher among Hispanics and lower among Blacks compared with Whites. The much lower inpatient open cholecystectomy rates were similar by sex and race-ethnicity. Ambulatory laparoscopic cholecystectomy rates were highest among middle-aged adults, followed by young adults and older adults. In contrast, both inpatient laparoscopic and open rates increased with age. For ambulatory laparoscopic cholecystectomy, insufficient years of data were available for trend analyses. From 2005 to 2019, inpatient cholecystectomy rates decreased overall by 30% for laparoscopic (112 to 78) and 60% for open (32 to 13) procedures and by sex, race-ethnicity, and age groups.

**Figure 4.**
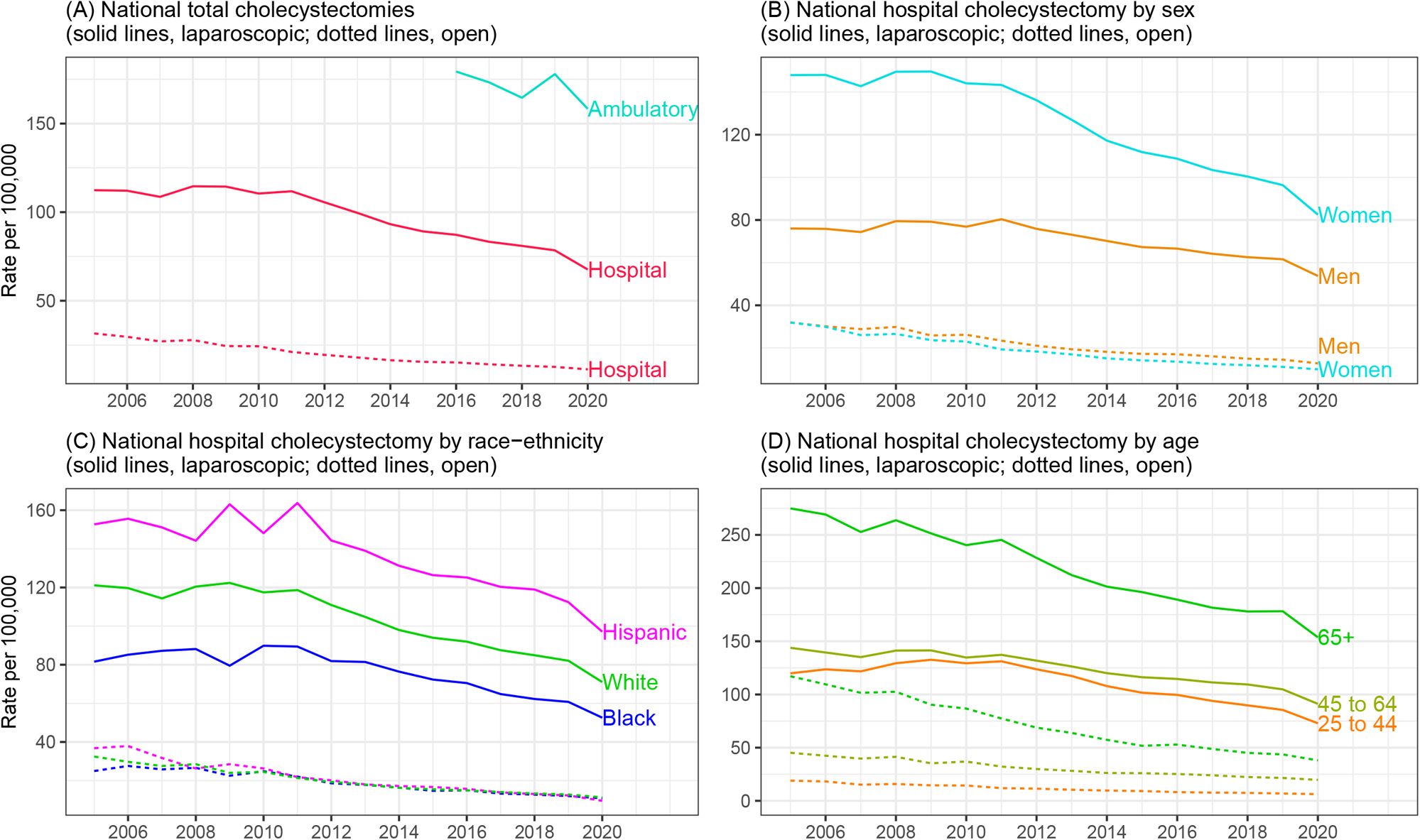
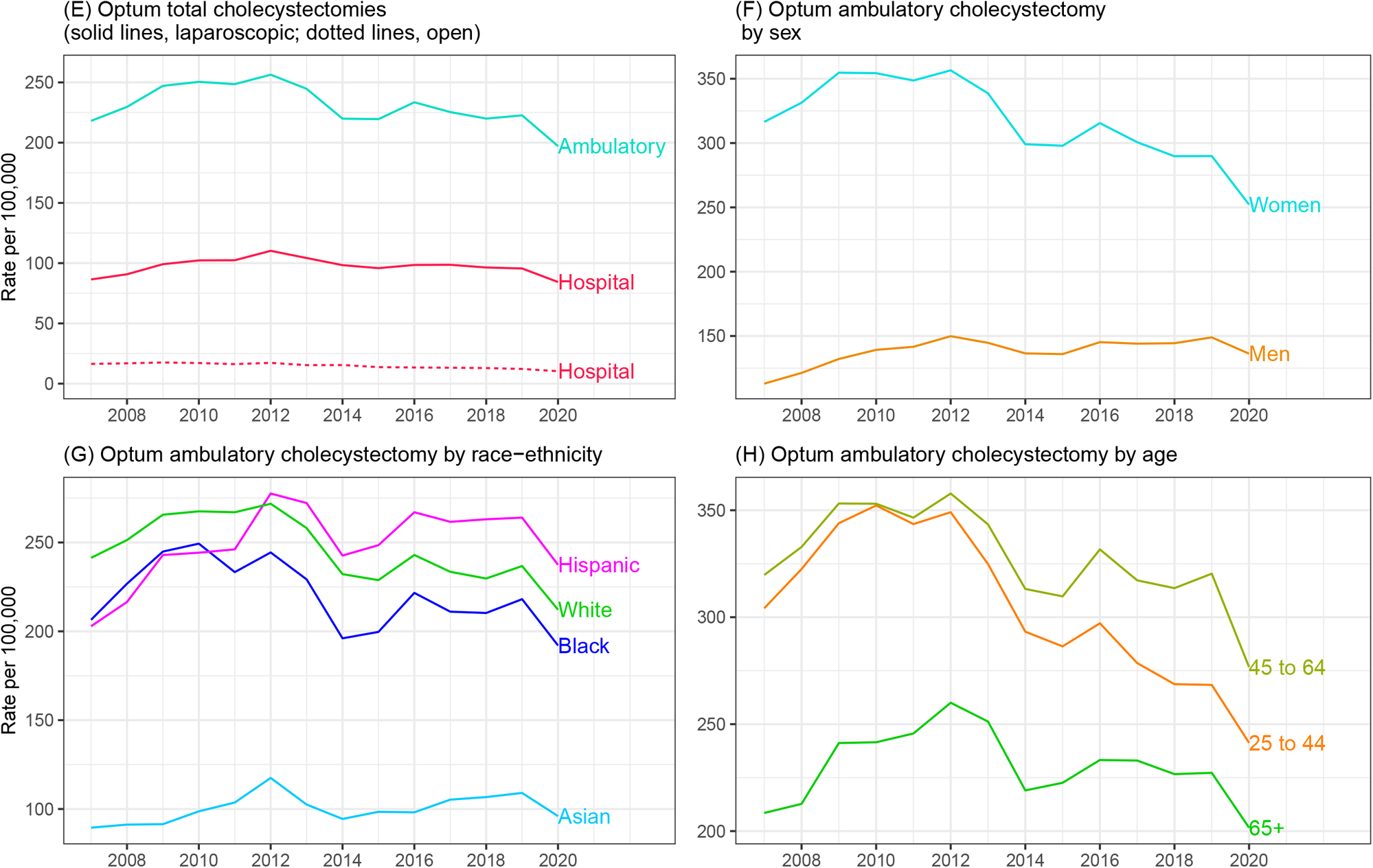
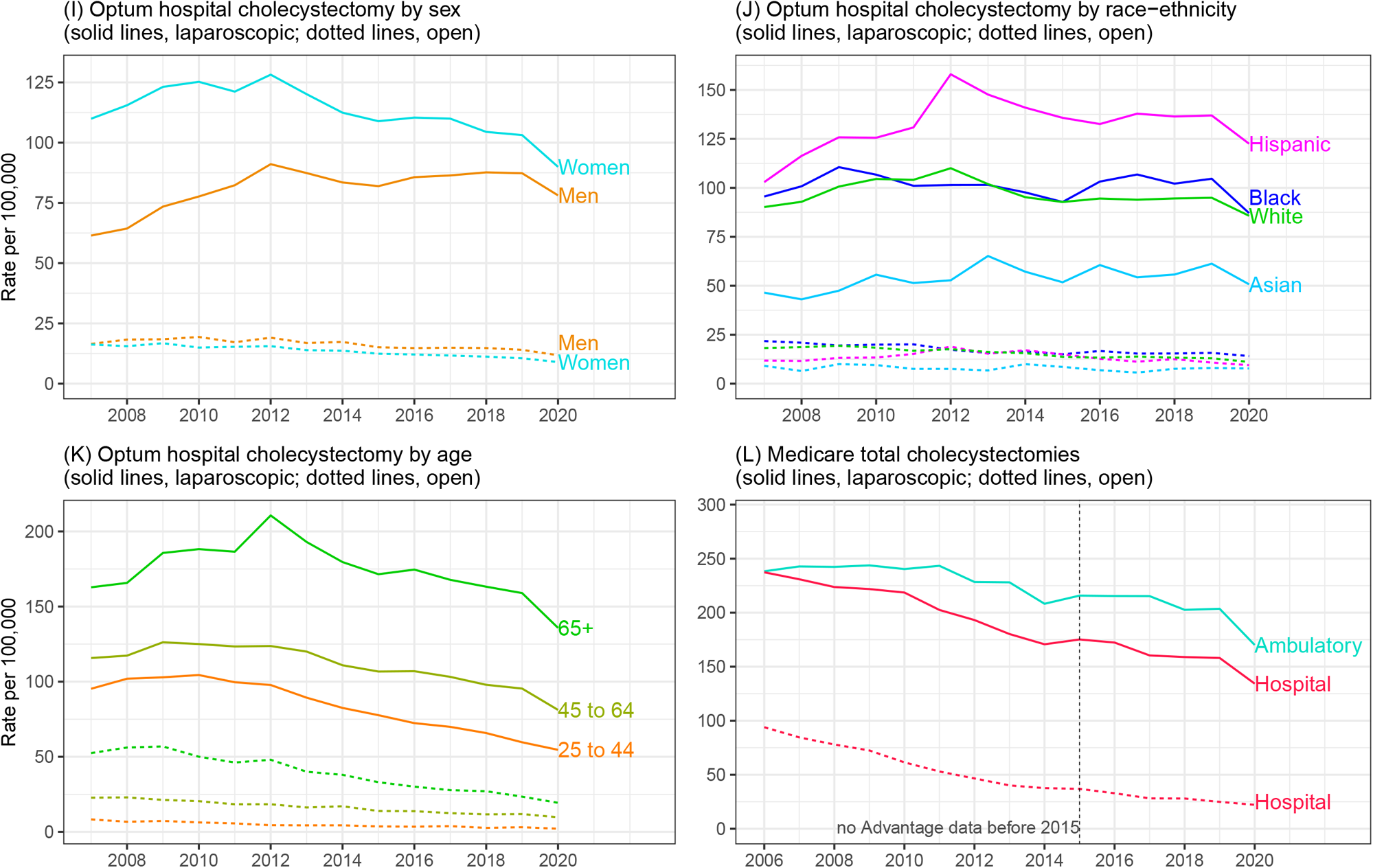
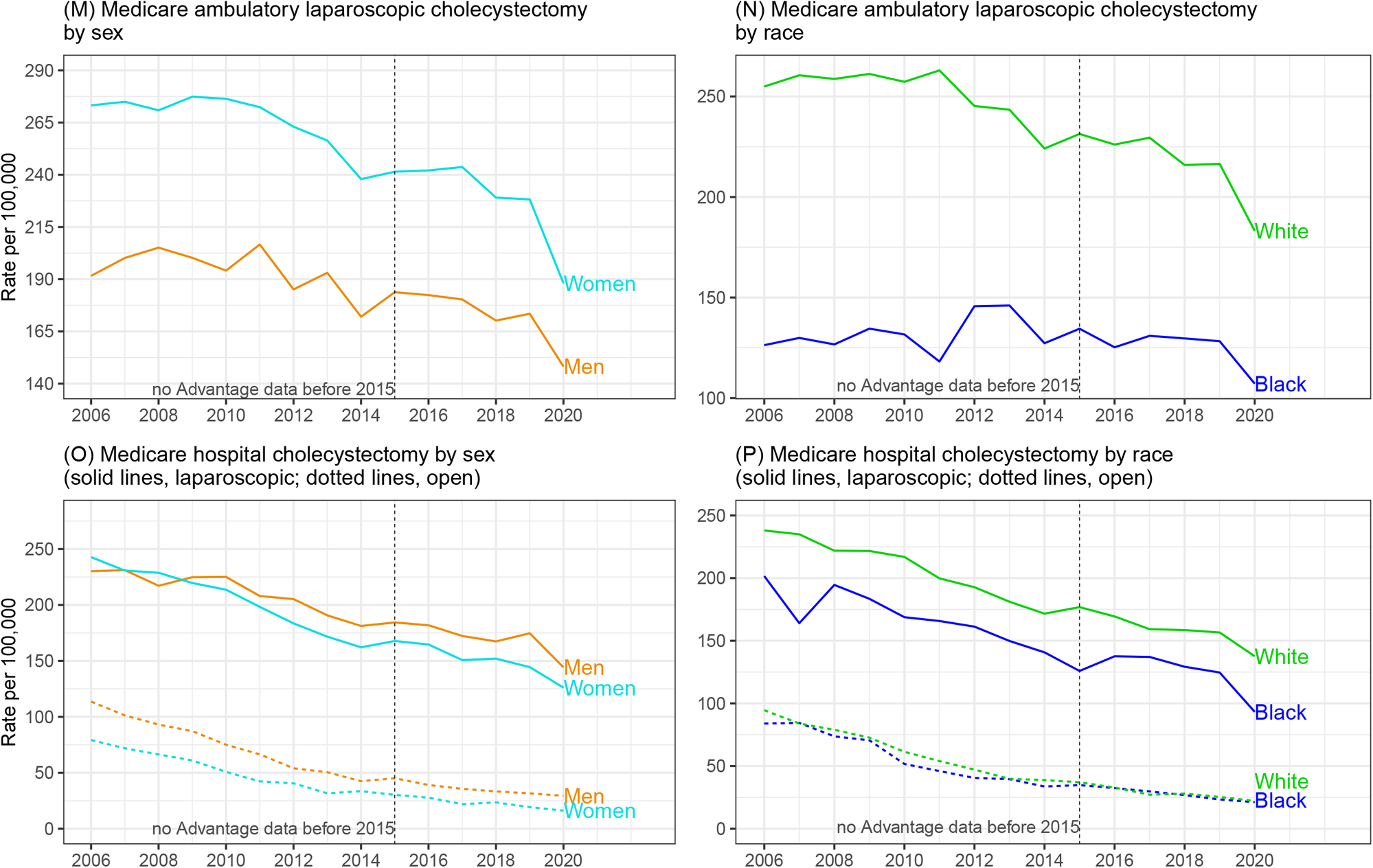
Cholecystectomy rates in the United States (Source: Healthcare Cost and Utilization Project (HCUP) Nationwide Ambulatory Surgery Sample, 2016-2020); HCUP National Inpatient Sample, 2006-2020; Optum Clinformatics® Data Mart, 2007-2020; CMS Medicare 5% file, 2006-2020) A. National total cholecystectomies B. National hospital cholecystectomy by sex C. National hospital cholecystectomy by race-ethnicity D. National hospital cholecystectomy by age E. Optum total cholecystectomies F. Optum ambulatory laparoscopic cholecystectomy by sex G. Optum ambulatory laparoscopic cholecystectomy by race-ethnicity H. Optum ambulatory laparoscopic cholecystectomy by age I. Optum hospital cholecystectomy by sex J. Optum hospital cholecystectomy by race-ethnicity K. Optum hospital cholecystectomy by age L. Medicare total cholecystectomies M. Medicare ambulatory laparoscopic cholecystectomy by sex N. Medicare ambulatory laparoscopic cholecystectomy by race O. Medicare hospital cholecystectomy by sex P. Medicare hospital cholecystectomy by race

### Cholecystectomy - commercial insurance data (Figures 4E-4K)

Among commercial insurance enrollees, rates (per 100,000 in 2019) were higher compared with national data for ambulatory laparoscopic (223 vs. 178) and inpatient laparoscopic (96 vs. 78) cholecystectomies, and similar for inpatient open (12 vs. 13) procedures. Among commercial insurance enrollees, laparoscopic cholecystectomy rates were highest among Hispanics and lowest among Asians. Ambulatory laparoscopic rates were lower among Blacks compared with Whites, while inpatient laparoscopic rates were similar. In contrast to national cholecystectomy rates that declined over the years studied, a decline was seen only for ambulatory laparoscopic procedures among commercial insurance enrollees, and only for women, Whites, and Blacks while rates increased for men and Hispanics.

### Cholecystectomy – Medicare data (Figures 4L-4P)

Among Medicare beneficiaries, rates (per 100,000 in 2019) were higher compared with national data for ambulatory laparoscopic (204 vs. 178) inpatient laparoscopic (158 vs. 78) and inpatient open (25 vs. 13) cholecystectomies. Like national data, ambulatory laparoscopic cholecystectomy rates were higher among female Medicare beneficiaries, but in contrast to national data, inpatient laparoscopic and open rates were higher among men. Among Medicare beneficiaries, laparoscopic cholecystectomy rates were lower among Blacks compared with Whites, especially for ambulatory procedures.

## DISCUSSION

In this report, we expanded on earlier findings and investigated current trends in the gallstone disease burden in the United States using national survey and claims databases.(15) The prevalence of gallstone disease has increased, especially among older adults, based on claims diagnoses among both commercial insurance and Medicare enrollees. Prevalence was higher among older adults, women, and Hispanics, and lower among Blacks (except among commercial insurance enrollees) and Asians, consistent with known risk factors. Medical care rates were higher among commercial insurance enrollees compared with national data for ambulatory care visits and hospitalizations, but lower for emergency department visits. Like gallstone disease prevalence, national ambulatory care visit, emergency department visit, and hospital discharge rates were higher among women. Among commercial insurance enrollees, higher rates among women compared with men were primarily seen for ambulatory care visits. In contrast to national data that included persons of all ages, among Medicare beneficiaries, emergency department visit and hospitalization rates were higher among men compared with women. National ambulatory care visit and hospital discharge rates were highest among Hispanics and ambulatory care visit, but not hospital discharge rates, were lower among Blacks compared with Whites. In contrast to national data, hospitalization rates were generally higher in Blacks among Medicare beneficiaries, as were emergency department visit rates. Medical care rates were lowest for Asians (among the commercially insured).

Rates of both ambulatory care visits to physicians’ offices and emergency department visits with gallstone disease increased since the mid-2000s. Hospital discharge rates declined beginning in the early 1990s due to the shift to outpatient laparoscopic cholecystectomy, then stabilized between 2000 and 2011, after which they again declined.(2) The decline in recent years was primarily among older adults (**Figure 1H**) and may be the result of two decades of widespread use of laparoscopic cholecystectomy. In contrast to national data, hospitalization rates increased among commercial insurance enrollees. Both ambulatory and emergency department visit rates increased since the mid-2000s.

Although the economic burden on the health care system from gallstone disease is high, the mortality rate associated with gallstone disease is low. Although gallstone disease is more common among women and women had higher medical care rates, the mortality rate was higher among men. In contrast to health care use which was higher among Hispanics compared with Whites, gallstone disease mortality rates differed little by race-ethnicity. Gallstone disease mortality has continued to decline in recent years resulting in an approximately 80% lower mortality rate over the past four decades,(3) primarily due to a decrease among older adults.

Most cholecystectomies are now performed laparoscopically in an ambulatory setting. Among commercial insurance enrollees, rates were higher compared with national data for laparoscopic cholecystectomies. National laparoscopic cholecystectomy rates were higher among women compared with men. In contrast, among Medicare beneficiaries, inpatient laparoscopic rates were higher among men. Among commercial insurance enrollees, laparoscopic cholecystectomy rates were highest among Hispanics and lowest among Asians. Ambulatory laparoscopic rates were higher among Whites compared with Blacks, while inpatient laparoscopic rates were similar. Ambulatory laparoscopic cholecystectomy rates were highest among middle-aged adults, followed by young adults and older adults, while inpatient laparoscopic and open rates increased with age. Among commercial insurance enrollees over the years studied, a decline was seen for ambulatory laparoscopic procedures, overall and among women, Whites, and Blacks, while rates increased among men and Hispanics.

Because of potential short-term effects of the COVID-19 pandemic on gallstone disease healthcare utilization and mortality, we calculated trends using rates for 2019, while figures present data through 2020. Between 2019 and 2020, there were generally small decreases in medical care use with gallstone disease and increases in gallstone disease mortality. Whether these changes represent only temporary fluctuations will become clearer when additional years of data are available.

The data used in this report have limitations. For national data sources, ambulatory care numbers and rates represent visits, not persons with a visit and hospital numbers and rates represent discharges, not persons with an inpatient stay. National health care data do not include care in federal facilities and NEDS and NASS include only hospital-owned emergency departments and ambulatory surgery facilities, respectively; therefore, rates based on the U.S. population are underestimates. The NAMCS sample is limited to office-based physicians, a group that has become a less inclusive source for ambulatory care, and samples are small so estimates are less statistically reliable. HCUP data sources do not include all states and not all participating states collect data on race-ethnicity. Mortality data are dependent on the accuracy of death certificates that may vary by condition and chronic diseases that contribute to mortality are frequently underreported.

The limitations are offset by the following strengths. NAMCS data were obtained from provider records. HCUP data sources are the largest all-payer inpatient, emergency department, and ambulatory surgery databases in the U.S. National health care and Medicare data were weighted to provide national estimates, and mortality data include all deaths occurring in the U.S. CDM and Medicare data are person-level and claim-based prevalence can be calculated. Medicare covers approximately 96 percent of all U.S. citizens aged 65 years and over permitting generalization to the U.S. older adult population.(14)

The burden of gallstone disease in the United States is considerable and rising. Although the gallstone disease mortality rate has decreased over the past four decades leading to a low case fatality rate, medical care devoted to gallstone disease is significant. Cholecystectomy is one of the most common digestive disease surgical procedures. Gallstone disease prevalence (claims based), ambulatory care visit, and emergency department visit rates are increasing, and the appearance of decreasing hospitalization rates is deceptive because of the shift from inpatient open cholecystectomy to ambulatory laparoscopic procedures. Significant disparities exist with a disproportionately high gallstone disease burden among women, Hispanics, and older adults. Current practice patterns should be monitored post-pandemic for better health care access. Public health measures are needed to address the substantial and increasing gallstone disease burden.

## Data Availability

Public use NAMCS data are available at https://www.cdc.gov/nchs/ahcd/index.htm. Vital Statistics of the U.S. Multiple Cause-of-death data are available at https://www.nber.org/research/data/mortality-data-vital-statistics-nchs-multiple-cause-death-data. Restricted use NAMCS data are not available according to NCHS guidelines. HCUP, Medicare, and Optum data are not available due to data use agreements.

https://www.nber.org/research/data/mortality-data-vital-statistics-nchs-multiple-cause-death-data

## ACKNOWLEDGMENTS

The authors thank Bryan Sayer, Helen Corns, Laura Fang, and Joe Evans for statistical programming.

